# Lived experience co-design of self-harm interventions: A scoping review

**DOI:** 10.1101/2023.08.18.23294271

**Authors:** Lucy C. Wright, Natalia Chemas, Claudia Cooper

**Author notes:** Corresponding author: Lucy C Wright, Barts and the London School of Medicine, Garrod Building, Turner St, London, E1 2AD, UK, 020 7882 5555.

## Abstract

**Background:** Self-harm prevalence is rising, yet service users encounter stigmatising attitudes and feel let down when they seek professional help. Co-design activities can potentially enable development of more acceptable and effective services.

**Objectives:** To map existing literature describing how people with lived experience of self-harm have engaged in co-designing self-harm interventions, understand barriers and facilitators to this engagement and how meaningfulness of co-design has been evaluated.

**Inclusion criteria:** Studies where individuals with lived experience of self-harm (first-hand or carer) have co-designed self-harm interventions.

**Methods:** In accordance with Joanna Briggs Institute (JBI) scoping review methodology we scoped PubMed, Embase, PsycINFO, Web of Science, Cochrane Library, PROSPERO, ClinicalTrials.gov and relevant websites on 24.12.22. A protocol was published online (http://dx.doi.org/10.17605/OSF.IO/P52UD). Results were screened at title and abstract level, then full-text level by two researchers independently. Pre-specified data was extracted, charted, and sorted into themes.

**Results:** We included twenty co-designed interventions across mobile health, educational settings, prisons, and emergency departments. Involvement varied from designing content to multi-stage involvement in planning, delivery, and dissemination. Included papers described the contribution of 110 female and 26 male co-designers. Few contributors identified as from a minoritized ethnic or LGBTQ+ group. Six studies evaluated how meaningfully people with lived experience were engaged in co-design: by documenting the impact of contributions on intervention design, or through post-design reflections. Barriers included difficulties recruiting inclusively, making time for meaningful engagement in stretched services, and safeguarding concerns for co-designers. Explicit processes for ensuring safety and wellbeing, flexible schedules, and adequate funding facilitated co-design.

**Conclusions:** To realise the potential of co-design to improve self-harm interventions, people with lived experience must be representative of those who use services. This requires processes that reassure potential contributors and referrers that co-designers will be safeguarded, remunerated, and their contributions used and valued.

**ARTICLE SUMMARY:** *Strengths and limitations of this study:*

1. Comprehensive search strategy with no restriction on publication date to capture breadth of evidence
2. All papers screened at title/abstract and full-text level by two researchers independently
3. Protocol uploaded to the Open Science Framework prior to conducting scoping review
4. Did not check all published self-harm intervention papers for evidence of co-design, so instances where co-design was not mentioned in the title or abstract could have been missed
5. Only the development paper for each intervention was included – follow up papers were excluded at full-text level which may have overlooked additional co-design details

## INTRODUCTION

As health services shift from paternalistic to person-centred care, there is increasing focus on engaging patients and carers with lived experience in designing services (1). Co-designed services are more efficient and relevant for end-users, foster positive emotions, and increase service-user knowledge (2). Gold standard co-design is both active and embedded, where those with lived experience are equal partners with a meaningful role incorporating creativity, problem-solving, and decision-making (3). Co-production comprises co-design alongside co-delivery (4, 5). Co-production guidelines state experiential knowledge should be respected by sharing power and decision-making, and building/maintaining relationships through continued dialogue and reflection. Ground rules should be established, reciprocity valued, and flexibility is crucial. Diverse perspectives should be sought, especially from underrepresented groups (6).

Involvement of experts-by-experience in mental healthcare design is widespread across early psychosis, eating disorders, adult psychological therapies, and youth mental health (7, 8). However, the state of the field of co-designed self-harm interventions has not to our knowledge been the topic of a published review.

Self-harm is defined as direct, deliberate harm to one’s own body in the absence of suicidal intent, for reasons not socially sanctioned (9). The most prevalent forms are cutting, burning, hitting, and banging (10). Self-harm is common. A nationally representative estimate of self-harm in England revealed a lifetime prevalence of 6.4%, with especially high rates in women aged 16-24, a quarter of whom self-harmed (11).

Self-harm is prevalent in patients with Complex Emotional Needs (12), with prevalence rates of 95% and 90% in adolescent and adult samples diagnosed with emotionally unstable personality disorder (13). Self-harm behaviour occurs across a wide range of psychiatric diagnoses. People with depression, substance use, and anxiety disorders are at particularly high risk (14). Self-harm is also present in the absence of comorbidities (15), prompting the inclusion of non-suicidal self-injury disorder as a condition in the DSM-V (16). High-risk groups include the LGBTQ+ population (17) and those with chronic physical illnesses (18). There are ethnic differences in self-harm presentation, with prevalence highest in Black females and repetition least likely in Black and South Asian individuals (19, 20).

Self-harm can serve to regulate distressing emotions and escape from negative internal states, communicate distress and self-punishment, and can serve an anti-suicide function for some (21, 22). However, self-harm is a strong risk factor for future non-suicidal self-harm and completed suicide, with suicide risk up to 49 times the general population (23, 24). All patients presenting with self-harm should receive information, have family/carers involved, undergo psychosocial assessment, and have a personalised care plan and risk assessment (25). A series of Cochrane reviews question the efficacy of existing psychological interventions. In children and adolescents, consistently positive outcomes were found for Dialectical Behaviour Therapy only (26), and in adults only Cognitive Behavioural Therapy-based psychotherapy and Mentalisation-Based therapy showed promise (27, 28).

Several streams of evidence suggest existing self-harm interventions are not fit for purpose. There are accounts of patients being refused pain relief in the emergency department due to the self-inflicted nature of their wounds - “I thought you liked pain” (29), or denied medical treatment under assumptions they would re-engage in self-harm (30). Patients recount stigmatising attitudes from healthcare professionals, labelled ‘attention-seeking’ for seeking help (31). Given the rise in self-harm in young people, it is particularly concerning that this age group report feeling let down by clinical services and dropped on discharge (32).

Patients’ perceptions are not unfounded. Clinical staff across emergency departments, general medical, and psychiatric settings had feelings of irritation and anger towards those presenting with self-harm (33). Unfortunately, these experiences are not unique to healthcare settings. Prison officers, nurses, and doctors reportedly exhibited hostility towards prisoners who engaged in self-harm (34).

Collaboration with patients and carers to design and implement new approaches and interventions may improve their acceptability and efficacy and build relationships with staff. While one systematic review noted that service-user evaluation of pre-designed psychosocial self-harm interventions was rare (35), there have been no attempts to synthesise research regarding whether and how people with lived experience have co-designed self-harm interventions. Given the stigma surrounding self-harm from medical professionals, as well self-stigma and the high number of people who self-harm who are not in contact with services (36), engaging this lived experience group may be particularly challenging. A review in this area is important to identify how co-design has been conducted and unique requirements and challenges to lived experience involvement.

### OBJECTIVES

The primary objective of this scoping review was to map the extent of lived experience involvement in co-designing self-harm interventions. We also sought to describe how representative co-designers have been of intervention end-users and explore benefits, challenges, barriers, and facilitators to co-design. Additionally, we aimed to examine how the meaningfulness of co-design has been evaluated. Scoping methodology afforded flexibility to identify and map key concepts and gaps in the literature (37).

## METHODS

This work followed the Joanna Briggs Institute (JBI) methodology for scoping reviews (38). The protocol is published (http://dx.doi.org/10.17605/OSF.IO/P52UD, Supplementary Material 1). Patients or the public were not involved in conducting this review.

### Eligibility criteria

We included studies where individuals with first-hand or carer experience of self-harm co-designed interventions, materials, or guidelines for self-harm. There were no restrictions on age, gender, diagnosis, or publication date. Primary studies, systematic reviews, meta-analyses, and grey literature were included. Only English language studies were included. Various collaborative design concepts such as co-design, co-production, co-creation, and Patient and Public Involvement were incorporated. Interventions solely for self-harm where the intention was to die were excluded. Involvement restricted to consultation or giving feedback on pre-existing interventions was excluded.

### Information sources

On 24th December 2022 we searched the following databases using a comprehensive search strategy comprising three concepts (‘co-design’, ‘self-harm’, and ‘intervention’): PubMed, Embase, PsycINFO, Web of Science and Cochrane Library, as well as grey literature in PROSPERO, and ClinicalTrials.gov. Supplementary Materials 2 and 3 detail the complete search strategy and example search. Websites were also scoped for relevant content: Department of Health, National Institute for Health and Care Research [NIHR], National Institute for Health and Care Excellence [NICE], The McPin Foundation, Royal College of Psychiatrists, Harmless, YoungMinds, MQ Mental Health Research and Mind.

### Procedures for analysis

Following de-duplication, all records were screened for eligibility at title and abstract level, then at full-text level, by two researchers independently (LCW and NC). Disagreements were resolved via discussion with a third person (CC). Decisions were recorded using Rayyan (www.rayyan.ai/).

LCW extracted the following data from included articles: authors, year and publication type, country, setting, intervention, self-harm definition, aims, methods, population, extent of co-design involvement, benefits/challenges, facilitators/barriers, remuneration. Pre-, during, and post-co-design activities were evaluated against key principles in NIHR guidelines for co-produced work including sharing power, including all perspectives, valuing contributor knowledge, reciprocity, and building and maintaining relationships (6). In line with scoping review guidelines, we did not formally assess the quality of included papers (39).

## RESULTS

Database searching returned 2737 records. Following deduplication, 1814 title and abstracts were screened for eligibility. 71 full texts were assessed, of which 17 were included. Two additional materials were identified through web searches and one through references. A final 20 studies were included. The PRISMA flowchart in Figure 1 summarises the selection process.

**Figure 1.**
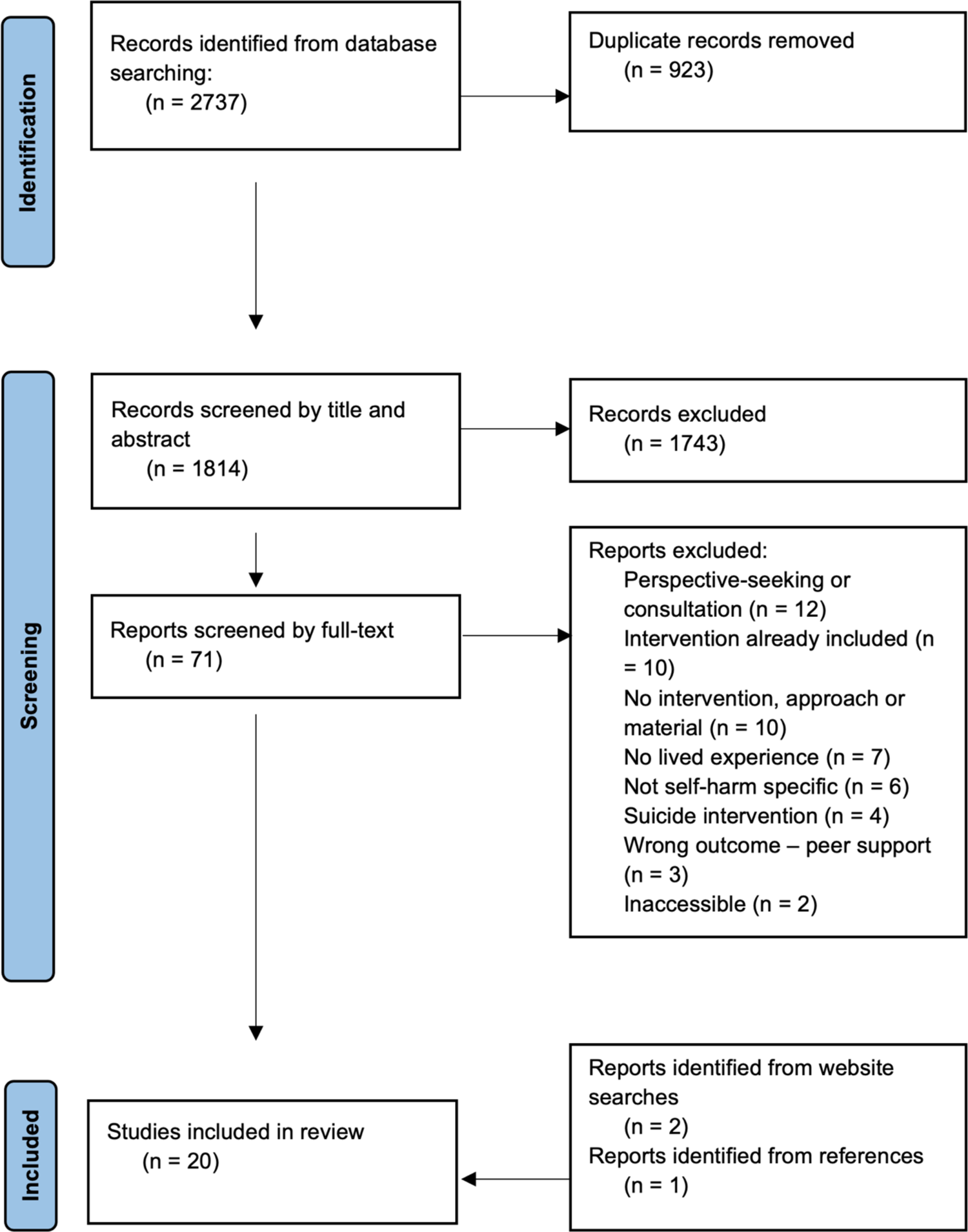
PRISMA flowchart of the screening process

Details of co-designed interventions, materials, and guidelines are outlined separately for young people (40–49) (Table 1) and adults (50–59) (Table 2). We included 13 qualitative studies, one quantitative, one mixed methods, one commentary, two protocols (for future co-design and evaluation of self-harm interventions), and two web pages outlining co-produced materials. Thirteen studies took place in the UK, the rest in Australia, India, USA, New Zealand, Canada, and Taiwan. All were published between 2005 and 2022.

Four interventions were for non-suicidal self-harm (40, 46, 52, 59), while six interventions did not discriminate based on suicidal intent (43, 44, 45, 47, 49, 58). The remaining sources did not define self-harm or the definition did not reference intent.

**Table 1.** Description of included studies involving young people in co-design of self-harm interventions.

| Author/year /country | Intervention / guideline / resource | Intended setting | Co-design aims | Methods | Stakeholders | Extent of involvement |
| --- | --- | --- | --- | --- | --- | --- |
| Bailey et al., (2019). UK (40) | Self-help materials for young people | Primary care | Develop materials to aid general practice self-harm consultations | Participatory action research: focus groups with young people, general practitioners, practice nurses; thematic analysis on focus group transcripts | N=15 aged 16-25 years with experience of self-harm recruited by snowball sampling<br>1 mixed race, 14 white; 8 M, 7 F<br>14 GPs and 16 practise nurses | Review and create materials, share help-seeking experiences |
| Latif et al., (2017). UK (41) | Our Care through Our Eyes e-learning for rCNs | General hospital | Co-produce a digital educational programme for rCNs caring for CYP who self-harm | rCN workshop: set priorities for their learning<br>CYP workshop: generate ideas for e-learning | 4 CYP admitted to CAMHS for self-harm treatment in past 12 months<br>Mean age 15 years. All F<br>19 nurses | Development workshop – shared hospital experiences, used flip charts and story boards to design training package, suggested how e-learning could be more engaging |
| Hetrick et al., (2018). Australia (42) | Symptom monitoring app ‘Wellbeing tracker’ Safety features | App for those in face-to-face treatment | Co-design app for young people with major depression/at risk of self-harm to self-monitor mood | 4 co-design workshops with young people<br>2 co-design workshops with clinicians – identify concerns | Current/former mental health service clients, treated for depression which may have included self-harm<br>Gender: 3 M, 8 F; Mean age 21.4 years<br>16 clinicians | Designed app features, individually and as group<br>1 young person presented design to clinicians<br>App designers sorted focus group notes into themes |
| Owens and Charles (2016). UK (43) | TeenTEXT: self-management text-messaging intervention | App for adolescents under CAMHS clinician | Re-develop adult text-messaging intervention for adolescents | Re-development: tailored components of adult intervention<br>Adapt intervention through its use<br>Focus group with CAMHS team – understand barriers to implementation | Development work: CAMHS patients with experience of self-harm, 3 clinicians<br>Feasibility work: 1 clinician-client dyad | Development work: creative workshops to alter adult intervention, delivery design by researchers, software developers, clinicians<br>Feasibility work: use intervention for 6 months and provide feedback |
| Stallard et al., (2018). UK (44) | Blueice smartphone app: toolbox of CBT/DBT strategies | App for 12–17-year-olds in face-to-face therapy | Create, refine, evaluate smartphone app<br>Evaluate Blueice | Co-production between young people, clinical staff, academics, app developers<br>Workshops with clinicians<br>Phase 1 evaluation of acceptability, safety | Development: young people with lived experience of self-harm and clinicians (n unknown, recruitment source unknown)<br>Evaluation: 40 young people with current/past self-harm | Co-produced app – design, layout, flow, content |
| Aggarwal et al., (2021). India (45) | ATMAN: counsellor delivered psychological intervention | Youth in LMIC | Co-design a scalable psychological intervention to reduce self-harm and improve functioning | Phenomenological thematic analysis of interviews; Systematic review - identify effective elements of existing interventions<br>2 rounds of development workshops | Youth aged 15-24 who presented to psychiatry department after self-harm (n=15); caregivers (n=4)<br>Development work: N=6 young people, N=5 MHPs health professionals (MHPs)<br>Finalising structure: N=7 young people, N=5 MHPs | Interviews on self-harm experience – identify intervention outcomes<br>Workshop round 1 - reflect on experiences, feedback on review, identify missing elements<br>Workshop round 2 – feedback to finalise structure |
| Kokaliari & Lanzano (2005). USA (46) | Consumer-therapist co-run self-injury group | American colleges | Design and run group for self-injury on principles of consumer empowerment | Plan and facilitate group | Members of consumer-run, campus mental health organisation with experience of self-injury<br>2 student consumers, 2 counsellors | Brainstormed solutions to increased self-injury<br>Planned group, delivered with professionals<br>Equal team members, all decisions made jointly |
| Meinhardt et al., (2022). New Zealand (47) | New Zealand specific guidelines for school staff managing self-harm | New Zealand high schools (age 12-19 years) | Develop culturally responsive guidelines for school staff supporting students who self-harm, consistent with bi-cultural principles | Delphi consensus method:<br>Literature review<br>Interviews with school staff<br>Expert panels (youth and stakeholders) chose items for guidelines<br>Rōpū Mātanga Māori resolved discrepancies | N=30 youth: aged >16 (2 no experience, 15 lived experience, 22 knew someone, 19 supported someone; 24 F, 5 M, 1 trans woman) recruited from youth advisory group<br>N=34 researchers, MHPs, school staff (1 no experience, 6 lived experience, 26 know someone, 28 supported someone)<br>46.9% European, 28.1% Māori, 18.8% Pacific Peoples, 6.3% Asian | Questionnaires – suggest new recommendations<br>Voted on which recommendations included in guideline<br>Acknowledged as guideline contributors |
| Bush (2016). UK (48) | No Harm Done: digital information pack, 3 short films | Variety | Co-create resources for people supporting those who self-harm | Collaboration between YoungMinds, The Charlie Walker Memorial Trust, Royal College of Psychiatrists | Young people, parents | Shared their stories |
| Knowles et al., (2022). New Zealand & UK (49) | Outcomes for Cochrane review of self-harm interventions for educational settings | Educational settings | Identify methods to generate outcomes, design outcomes important to young people, compare to typical review outcomes | Participatory co-design workshops<br>Thematic analysis on generated ideas | N=28 young people with individual or friend/family experience of self-harm<br>New Zealand: 5 M, 8 F between 16-23 years: 6 Māori, 4 New Zealand Europeans, 4 Asian<br>UK: 11 F, 3 M, 1 non-binary<br>Recruited from professional networks and young people’s organisations | Workshops: design review and generate outcomes<br>Pick 3 ‘must include’ and ‘must exclude’ outcomes<br>Collaboration to reach final 6 outcome themes<br>Input on how outcomes could be measured |

GP = general practice; CYP = children and young people; rCN = registered children’s nurses; CAMHS = child and adolescent mental health services; CBT = cognitive behavioural therapy; DBT = dialectical behavioural therapy; LMIC = low- or middle-income country; MHPs = mental health professionals; F = female; M = male

**Table 2.**
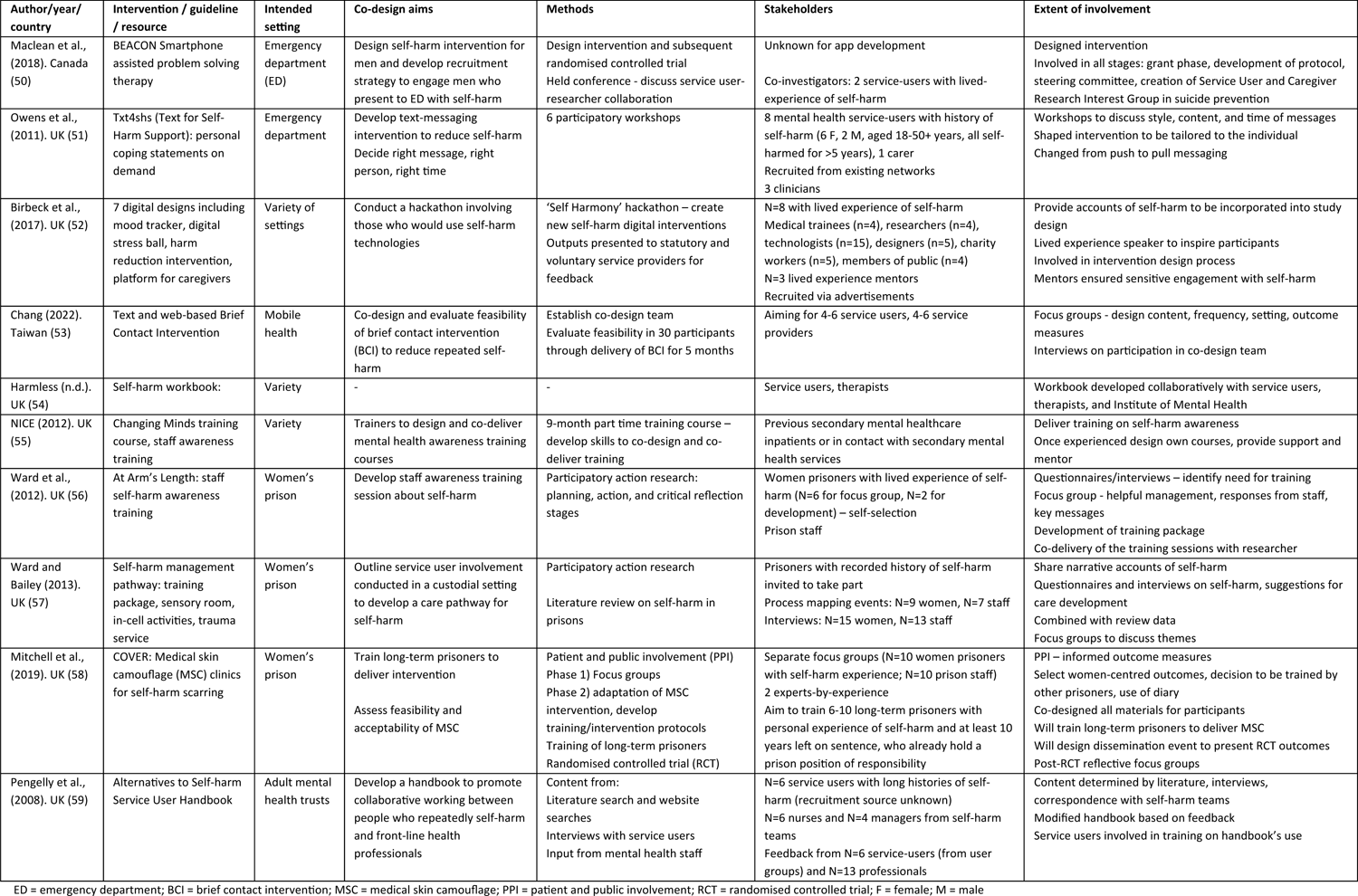
Description of included studies involving adults in co-design of self-harm interventions.

### What interventions have been co-designed?

Of the 10 interventions designed by and for children and young people, three were mobile health technologies for use between face-to-face sessions with mental health professionals (42–44). Four were resources to support care providers – primary care practitioners (40), general hospital children’s nurses (41), parents and teachers (48), and school staff (47). In educational settings, young people co-designed and co-ran a self-injury group (46) and co-designed outcomes for self-harm interventions which informed a Cochrane review (49). Youth in India co-designed a psychological intervention for use in low and middle-income countries (45).

Mobile health interventions were also the focus of three interventions co-designed by adults with lived experience of self-harm (50–52), and one protocol for a planned brief contact intervention (53). Adults also co-designed materials to aid professionals and carers, including a handbook for mental health trusts (59) and self-harm awareness training delivered by experts-by-experience (55). Women’s prisons were the focus of three interventions (56, 57, 58). Finally, adults co-designed an activity workbook for self-harm recovery (54).

### To what extent were individuals with lived experience involved?

#### Before co-design

Four studies describe how people with lived experience were involved in planning how studies would be conducted or evaluated, or in securing funding. In one study, people with lived experience identified the need for a co-run self-injury group within their American college campus (46). Service-users were also involved at the grant phase and protocol development (50), and informed study outcomes (58).

#### During co-design

One study engaged people in online workshops (49) while the remaining co-design was face-to-face via workshops, focus groups, and interviews. Using sticky notes, co-designers wrote and thematically sorted triggers, urge-reduction messages, and characteristics of groups who self-harm for Txt4shs, with further workshops to personalise and refine the intervention (51). Over four workshops young people sketched intervention features as individuals, obtained group feedback, then prioritised optimal features for the final group design (42).

Using information gleaned from other stakeholders or literature alongside service-user design was common (59). Themes emerging from statistical analysis of medical records and challenges identified by general practice staff guided focus groups to source and create self-help materials for self-harm consultations (40). Registered children’s nurses identified their training needs, upon which workshops were held with children and young people who used storyboards to reflect on their experiences and decide what should be included (41).

Voting was frequently used in decision-making. Designing a psychological intervention, youth added missing elements to those identified through interviews and a systematic review, voted on elements for inclusion and built elements into modules (45). Young people co-designed a review for self-harm interventions by anonymously suggesting review outcomes which were combined with typical outcomes recorded in trials and voted on for inclusion (49). Using the Delphi consensus method, stakeholders voted on recommendations obtained from literature searches and interviews with professionals and experts-by-experience for inclusion in school self-harm management guidelines (47).

The Self Harmony hackathon uniquely included people with lived experience as designers, as inspiration through sharing their experiences, and as mentors to ensure sensitive engagement with self-harm (52). *No Harm Done* materials were also unique since sharing self-harm stories on film was the co-creation contribution, using real-life experiences to dispel myths (48).

Extent of co-design involvement was less clear when interventions were not afforded a separate development paper (50) - creative workshops (43), creating, refining, and evaluating an app (44), and collaborative development of a prison self-harm pathway (53) and self-harm workbook (54). Service-users will be involved in developing the content, settings, and outcomes of a brief contact intervention (53).

Four studies involved co-delivery. Students planned topics for and facilitated a college self-injury group alongside counsellors (46). People with lived experience also designed and co-delivered self-harm awareness training (55). In prisons, women designed outcomes for an existing intervention (58) and a staff training package (56) which will be delivered by other prisoners with self-harm experience.

All decisions regarding the co-run self-injury group were made between consumers and counsellors who were viewed as equals (46). However, elements of some interventions were determined prior to lived experience involvement - content type and web-based nature (53), mood monitoring features (42), and an existing intervention for re-development (43).

#### After co-design

App design ideas (42), handbook training (59), and findings (58) were co-disseminated. Only one paper explicitly stated those with lived experience were acknowledged as contributors on final guidelines (47). Continued dialogue was rare, though people with lived experience not only co-designed the BEACON intervention but were co-investigators in a subsequent randomised controlled trial (50).

#### Remuneration

This varied from a certificate (47) to travel reimbursement and food provision (52), vouchers (41, 47, 49), hourly pay (42, 51) and unspecified payment for service delivery (55). More attractive incentives were proposed to encourage recruitment (41). Prison settings did not detail reimbursement, but stated the intervention would not interfere with women’s income (58). Others offered training opportunities such as conference attendance (50). No papers outlined the rationale for their chosen reimbursement, nor the time commitment of contributors.

### Who is involved in co-design?

Most work included individuals with personal self-harm experience recruited via services (41, 45), existing team networks (42, 47, 51), young people’s organisations (48, 49), advertisements (52), or college mental health organisations (46). Snowball sampling was common (40, 42, 50). To manage risk, some studies excluded individuals who self-harmed in the past three months (42) or were receiving acute hospital care for their self-harm (41). There was some gatekeeping to involvement by healthcare professionals and prison staff who excluded people if they were not deemed suitable for workshops (41) and selected prisoners who were most ‘suitable’ for intervention delivery or already held positions of responsibility (56, 58). Five studies also involved parents or carers (45, 48, 49, 47, 51). In some studies, co-designers varied across the development process (51) or new individuals were added to make final modifications (45).

In studies reporting demographics of lived experience co-designers, 110 were female and 26 were male (40, 41, 42, 47, 49, 51, 56, 57, 58). One non-binary person and one trans woman were also included (47, 49). Few studies reported ethnicity. Young people who co-designed materials for UK general practice were overwhelmingly white (40), while New Zealand studies sought Māori and non-Māori representation and recruited a Rōpū Mātanga Māori (clinical cultural governance group) to ensure Māori-centred work, given higher self-harm rates among this population (47, 49). A study based in India recruited from the local population to develop an intervention for low- and middle-income countries (45). No studies presented information on sexual orientation, self-harm frequency (besides meeting an inclusion cut-off), or comorbidities.

### Was co-design meaningful?

Meaningfulness of lived experience involvement may be discerned from how co-design benefited the intervention or reports from co-designers on the impact of their involvement. Several papers outlined positive impacts of their co-design efforts but did not report how these were assessed, for example enabling the lived experience voice to be heard (41, 42, 46, 49) and making interventions relevant to end-users (41, 47, 50). Co-deliverers reportedly broke down barriers to professional-run groups, served as role models for attendees (46), developed transferable skills (55), provided meaningful work, and addressed the inmate-officer divide of a prison setting (57, 58). However, few studies quantified the degree or success of these activities.

Three studies explicitly documented how lived experience contributions impacted intervention design. Young people identified more asset-based outcomes for self-harm interventions (‘better coping’ and ‘safer environment to talk about self-harm’) than typical self-harm reduction/cessation, prompting researchers to transform their review (49). Researchers were challenged on their preconceived idea to sub-categorise people who self-harm and send generic support messages at pre-specified times. Highlighting the personal nature of self-harm and potentially detrimental effects of receiving blanket messages paved the way for the highly personalised Txt4shs app (51). The Self Harmony hackathon informed a platform where digital mental health tools will be open-sourced (52).

Three studies involved reflections on the co-design process. Assessments conducted with Changing Minds co-trainers revealed involvement gave them a valued role, increased self-esteem, and confidence to develop supportive social networks and challenge discrimination (55). One co-deliverer of prison self-harm awareness training reflected how the experience increased their self-esteem, confidence, and acceptance of their own self-harming frequency. Additionally, most staff recipients reflected that the lived experience perspective was the most useful element (56). Reflective focus groups with young people and clinicians highlighted short consultations as a limiting factor of their co-designed materials (40). Some studies conducted debriefing but did not include what was discussed (46, 49).

### What were the barriers and facilitators to self-harm intervention co-design?

#### Recruiting people with lived experience

Recruitment challenges precluded co-design (41), and co-designers were only included if deemed suitable by parents/guardians, clinical, or prison staff (41, 56, 58). Attrition was high due to individuals being too unwell (43), fluctuations in mental state and personal circumstances (51), and study length (47). Support to withdraw and re-join (50), more incentives, and online opportunities were suggested strategies to encourage involvement (41).

#### Safeguarding

Feared repercussions by people with lived experience may have prohibited their involvement, for example peers becoming aware of their self-harm history (42), relationship issues between prisoners and staff (56), and self-harm exacerbation (57). Indeed, an individual with self-harm lived experience reported that co-running a self-harm group was intense and draining (46). Researchers reported various measures to mitigate for some of these concerns (41) such as ground rules regarding personal disclosure (45, 49), completion of a “wellness plan” (not elaborated on) prior to participation (42), support from youth workers and clinical psychologists (49), mental health charity input to ensure co-design activities are sensitive, and provision of safe spaces to relax or obtain support from volunteers (52). Studies used follow-up debriefing and phone-calls to check wellbeing (46, 49).

#### Enabling collaborative involvement

Too didactic or formal methods with minutes and agendas were difficult to engage with (49, 50). A ‘persona’ method was used to overcome challenges eliciting review outcomes whereby cases of young people self-harming were presented, and co-designers were asked how cases would be better after a successful intervention (49). Skills deficits, for example in scientific literature searching, limited lived experience involvement (51). Power imbalances, particularly within the prison system, were a barrier to collaborative involvement (57). Facilitatory measures included creating a safe space (50) and assuring there were no right or wrong answers (41). Placing service-users in leadership roles also avoids tokenistic involvement (55, 57). Ensuring co-design was a two-way relationship was an additional facilitator, with service-users benefitting from skill development (42), training opportunities (50) and payment (55).

#### Time and funding

Radical revision of predetermined ideas slowed development of a co-designed app and required flexibility, only possible with funding bodies willing to tolerate uncertainty (51). Adequate funding is needed to build relationships with clinical teams and cover the timespan necessary to incorporate lived experience input (43, 55).

#### Wider mental health system

Service-user designs must be considered against clinician availability (42) and professional views (59). TeenTEXT was unable to go through further co-development as new technology burdened burnt out Child and Adolescent Mental Health Services (43). Institutional stigma regarding the capacity of service-users to deliver training was a barrier to co-delivery (55). Presence of professionals facilitated co-production, supporting those who may otherwise have been reluctant to run a self-injury group (46). Barriers like service capacity should be anticipated and rectified early (43), and new practices are required rather than trying to fit co-design into typical research practices (50).

## DISCUSSION

In this scoping review we identified twenty co-designed interventions, approaches, and materials for self-harm across settings. Though co-design arose in the 1970s (60), most studies were published in the 2010s, in the UK. This surge in co-design publications is perhaps unsurprising given increasing self-harm prevalence particularly in young people and the recent push towards lived experience involvement (61, 62). Ten interventions were designed by and for children and young people and ten by adults. Where characteristics were reported, co-designers were predominantly women and were in contact with mental health or prison services. This was the first review to explore depth of lived experience involvement in the self-harm field, factors that help and hinder co-design, and meaningfulness of involvement. A robust search strategy across multiple databases enabled thorough examination of the literature.

Our findings indicate lived experience co-design varied from designing aspects of interventions with considerable input from the literature and other stakeholders, through to multi-stage involvement in design, delivery, and dissemination, with equal decision-making say. It may be misinformed to aim for equal involvement in all decisions - guidelines state there can still be a leader, whether they are a service-user or another stakeholder (6). Few studies fostered involvement beyond initial design activities which may be viewed as tokenistic if co-designers are unable to see the impact of their involvement, particularly having shared personal information (63, 64).

Many stated benefits of co-design such as making interventions relevant to end-users and breaking down the staff—service-user divide lacked tangible empirical or qualitative evidence. Barriers and facilitators of co-design fell into themes of recruitment, safeguarding, involvement methods, time and funding, and mental health services. Meaningful co-production should be “equitably remunerated” (65) and “commensurate with the nature and demands of the activity” (66) though, where reported, remuneration varied from a certificate of participation to hourly pay and did not meet recommendations (67, 68). More transparency is therefore required in the compensation process.

Unrepresentative stakeholders, or involvement activities that exclude the most vulnerable in society could perpetuate power imbalances in self-harm interventions. Co-designers were predominantly cis-gender women, especially in prison systems where co-design only took place in women’s institutions. While this gender imbalance reflects self-harm prevalence, a significant number of men are affected (69, 70). Ethnicity was infrequently reported. Higher-risk groups including those acutely unwell, the LGBTQ+ population (71), and those with physical or mental health comorbidities were underrepresented. Self-harm may present differently in the context of certain conditions and tailored interventions may be required. Additionally, since self-harm is a somewhat hidden phenomenon, interventions designed by those in contact with services may not represent needs of the wider population who self-harm (62). While online workshops remove geographical constraints to participation, they may be prohibitive for those lacking technology access. Indeed, research suggests experts-by-experience should be provided with the necessary equipment to remove barriers to involvement (64).

Strategies such as snowball sampling and recruitment via existing networks may explain the lack of diversity in these lived experience samples. There was an element of clinician gatekeeping such that only those deemed suitable to take part acted as co-designers, though the criteria for suitability was often not reported. It is conceivable that ethics committees may have prohibited involvement of those at greatest risk to themselves, but greater transparency documenting the inclusion process is required to confirm this.

### Limitations

Though our search strategy was comprehensive across multiple databases, papers where search terms were not referenced in the title or abstract may have been overlooked. Our exploration of the representativeness of co-designers was limited by several papers not describing characteristics of those involved. Given the imperative for co-design of services in many countries, the relative paucity of evidence found suggests many co-design activities may be unpublished.

## Conclusions

Co-design of self-harm interventions is becoming more frequent, but work is required to improve representation from ethnically diverse, male, and higher-risk individuals. Additional safeguarding measures and support from relevant mental health or LGBTQ+ champions to ensure sensitive involvement could empower a wider group to have their voices heard. Conceivable financial, technological, and systemic barriers must be broken down and awareness raised of co-design opportunities to increase accessibility. Importantly, more transparency is required when documenting decisions surrounding the co-design process.

## Data Availability

Scoping review protocol, full search strategy, and example search strategy are available via the Open Science Framework.

http://dx.doi.org/10.17605/OSF.IO/P52UD

## ACKNOWLEDGEMENTS

This work was presented as a poster at the 7^th^ Suicide and Self-harm Early and Mid-Career Researchers’ Forum (EMCRF).

## FUNDING STATEMENT

This research received no specific grant from any funding agency in the public, commercial, or not-for-profit sectors.

## AUTHOR CONTRIBUTIONS

LCW and CC conceptualised the scoping review. LCW and NC read papers at title/abstract and full text level and decided on inclusion. LCW wrote the manuscript. All authors contributed to interpretation of results and editing the manuscript.

## COMPETING INTERESTS STATEMENT

All authors have completed the ICMJE uniform disclosure form at https://www.icmje.org/disclosure-of-interest/ and declare: no support from any organisation for the submitted work; no financial relationships with any organisations that might have an interest in the submitted work in the previous three years; no other relationships or activities that could appear to have influenced the submitted work.

## DATA SHARING

Scoping review protocol, full search strategy, and example search strategy are available via the Open Science Framework (http://dx.doi.org/10.17605/OSF.IO/P52UD).

## Notes

### Competing Interest Statement

The authors have declared no competing interest.

### Clinical Protocols

http://dx.doi.org/10.17605/OSF.IO/P52UD

### Funding Statement

This study did not receive any funding

