## Supplementary Materials for "Lived experience co-design of self-harm interventions: A scoping review"

**Supplementary material 1. Original protocol**

Co-design methods for self-harm interventions: a scoping reviewprotocol

What is co-design?

As health services have shifted away from paternalistic medicine to person-centred care "with, not for" the patient, there is increasing focus on what patients and carers with lived experience can bring to services (Sunkel and Sartor, 2022). Co-design refers to the collaborative involvement of end-users alongside other key stakeholders in the design of research, services and initiatives (Burkett, 2012). This approach recognises the unique knowledge held only by patients and carers with first-hand experience. Gold standard co-production is both active and embedded, where those with lived experience are equal partners who take a meaningful, active role including creativity, problem-solving, and decision-making (Rethink Mental Illness, 2017). In this way, co-design entails more than mere consultation or feedback provision, or choosing between options already decided by another party. Co-designed research and services are more efficient and relevant for end-users, foster positive emotions and increased knowledge for end users, and promote creativity within the organisation (Steen et al., 2011). In fact, within the research sphere, theDepartment of Health's 10 year strategy for mental health research recommends funders require user-led involvement, particularly increasing representation of children and young people and those with protected characteristics (Department of Health and Social Care, 2017). Involvement of experts by experience in mental healthcare is widespread, including early psychosis, adult psychological therapies, and youth mental health (Green et al., 2020). A review of eating disorder literature highlighted that co-production varies from sharing stories to providing guidance and designing interventions, through to peer-mentoring programmes (Lewis and Foye, 2021). Despite studies across other contexts, the state of the field of co-design in self-harm interventions is not known.

Self-harm:

Self-harm is defined as direct, deliberate bodily harm in the absence of suicidal intent, for reasons not socially sanctioned (such as religious rituals). The most prevalent forms are cutting, burning, hitting and banging, but it can also involve interfering with wound healing (Cipriano et al., 2017). Self-harm is common. A study of a nationally representative adult sample in England revealed a lifetime prevalence of 4.86%, while estimates in adolescents rise to 18% (Liu, 2021, Glenn and Nock, 2022). Self-harm is prevalent in patients with Complex Emotional Needs (CEN) - a descriptor for people likely to be diagnosed with "personality disorders" that recognises the controversy around such diagnoses (Foye et al., 2022). Self-harm is a key diagnostic criterion for emotionally unstable personality disorder with prevalence rates of 95% and 90% in adolescent and adult samples respectively (Goodman et al., 2017). However, self-harm behaviour is seen across a wide range of psychiatric diagnoses, with those with depression, substance use, and anxiety disorders at particularly high risk (Haw et al., 2001). Importantly, self-harm is also present in non-clinicalsamples (Swannell et al., 2014), prompting the inclusion of non-suicidal self-injury disorderas a condition in its own right in the DSM-V (Zetterqvist, 2015).

Self-harm can serve a range of functions to the individual. Intrapersonal functions, especiallyregulation of distressing emotion and escape from negative internal states are most commonly reported; other functions include communicating distress and self-punishment (Taylor et al., 2018). Counterintuitively, self-harm behaviour can actually serve an anti- suicide function (Klonsky, 2007). Self-harm is a strong risk factor for future non-suicidal self- harm and completed suicide, with risk up to 49 times the general population (Fox et al., 2015, Hawton et al., 2015). These findings, coupled with increasing rates of self-harm highlight the growing need for effective self-harm interventions (McManus et al., 2014).

Self-harm is managed across primary and secondary care, as well as in educational settings and the criminal justice system. NICE guidelines state that all patients presenting with self- harm should receive information, have family/carers involved, undergo psychosocial assessment and have a personalised care plan and risk assessment (National Institute for Health and Care Excellence [NICE], 2022). There are a range of psychological interventions available for self-harm, such as Dialectical Behaviour Therapy (DBT), Mentalisation-Based Therapy (MBT) and remote-contact interventions. Interestingly, recent advances in phone and web-based interventions that overcome barriers to face-to-face treatments have shownpromise in reducing non-suicidal self-injury (Arshad et al., 2020). However, a series of Cochrane reviews call the efficacy of such interventions into question. In children and adolescents, consistently positive outcomes were found for DBT only (Witt et al., 2021a). In adults, Cognitive Behavioural Therapy-based psychotherapy and MBT showed promise but evidence for other interventions including medication was uncertain (Witt et al., 2021b, Witt et al., 2021c). Thus, existing self-harm interventions differ in their efficacy across groups.

Several streams of evidence suggest that existing self-harm interventions are not acceptableto their users. The emergency department is a particular context in which many patients report negative experiences of interventions. Patients and carers perceive there to be a lackof parity of esteem between self-harm and physical healthcare interventions. For example, there are accounts of patients being refused pain relief due to the self-inflicted nature of their wounds - “I thought you liked pain” (DrEm_79, 2016), or denied medical treatment under the assumption they would likely re-engage in self-harm behaviour (Quinlivan et al., 2021). Moreover, some patients report a lack of sympathy from staff or humiliation during physical treatment for their injuries. Patients, particularly those with a personality disorder diagnosis, recount stigmatising attitudes from healthcare professionals, labelled as ‘attention-seeking’ for attending emergency departments for help following self-harm (Taylor et al., 2009). Patients’ perceptions are not unfounded. A systematic review revealed that clinical staff across emergency departments, general medical, and psychiatric settings had generally negative attitudes towards those presenting with self-harm - particularly feelings of irritation and anger (Saunders et al., 2012).

The problem is made worse by the wider system failing people who self-harm through longwaiting lists and exclusion of those with complex needs or without a diagnosis. People whoself-harm therefore have no choice but to access crisis support in the emergency department (O'Keeffe et al., 2021). Unfortunately, these experiences are not unique to healthcare settings. Prison officers, nurses, and doctors reportedly exhibited hostility towards prisoners who engaged in self-harm during their sentence (Marzano et al., 2012).

Given the rise in self-harm in young people, it is particularly concerning that this age group report feeling let down by clinical services. These experiences can be driven by limitations ofchild and adolescent mental health services at the organisational level such as waiting times, non-personalised therapies and the perception of being ‘dropped’ when discharged (Wadman et al., 2018b) but also at the individual level, feeling patronised by the healthcare professional they saw (Wadman et al., 2018a).

Why is a scoping review warranted?

Users’ needs are not being met by current interventions. Patient and carer suggestions to improve services include increasing patient decision-making, specific self-harm training for healthcare professionals, increasing sympathy from staff, improving access to immediate and post-self-harm care, and education to challenge stigmatising views (Taylor et al, 2009). Co-design with patients and carers to design and implement new approaches and interventions may improve the acceptability and efficacy of self-harm interventions and build relationships between staff, patients, and carers. It is not known whether existing co- design efforts are inclusive of those at increased risk of self-harm, for example the LGBTIQ population and those with chronic physical illnesses (Marchi et al., 2022, Singhal et al., 2014), nor whether co-design is representative of those who will ultimately use the service given some disparities in the nature and presentation of self-harm across ethnicities (Bhui etal., 2007). Furthermore, it is possible there would be challenges to its implementation. For example, involvement may bring back distressing emotions for some individuals so risk assessment and support during the process is critical (Lockwood et al., 2018).

The extent to which co-design is currently utilised in self-harm interventions is unclear. There have been no attempts to synthesise existing research to examine whether and howco-design has been put into practise and identify the co-design benefits and challenges specific to self-harm.

Aims and Objectives:

The aim of this scoping review is to identify and map available literature on how people withlived experience of self-harm are involved in co-designing interventions for self-harm. More specifically, our objectives are as follows:

1. Explore the extent of involvement of people with lived experience of self-harm in

co-designing interventions by examining the depth of involvement and what stages of the process they collaborated on
2. Identify the settings that interventions have been co-designed to take place in (e.g.primary or secondary care, digitally)
3. Explore who is involved in the co-design process - does this reflect the demographicof individuals who self-harm? Are co-design methods inclusive?
4. Highlight barriers and facilitators to co-designing self-harm interventions
5. Highlight benefits and costs of co-design of self-harm interventions
6. Identify any gaps in the co-design literature for self-harm interventions

A scoping review is the most appropriate method to investigate these objectives as it enables description and mapping of key concepts within the field rather than assessing study quality or performing quantitative synthesis. Scoping affords greater flexibility than asystematic review as studies with varying methodologies will be incorporated, enabling an exploration of the full extent of the available literature along with any gaps (Peterson et al.,2017, Munn et al., 2018).

To our knowledge, there has been no published research on this topic. A protocol for a scoping review of co-production across mental health service provision has been publishedand while self-harm interventions may be incorporated, its breadth would not permit our objectives to be met (Norton, 2022). The current scoping review will consider self-harm intervention co-design across clinical and non-clinical populations, as well as incorporating online and self-management interventions, which are beyond the scope of Norton's healthservice protocol.

### Methods

The proposed scoping review will follow the JBI methodology for scoping reviews (Peters etal., 2022). The protocol will be made publicly available via Open Science Framework (OSF, https://osf.io/). In line with guidelines, ethical approval is not required for this scoping review.

#### Eligibility criteria

Population: Literature included in this scoping review will involve participants with first- hand or carer experience of self-harm who have been involved in co-designing interventions, approaches, or informational material for self-harm. There will be no restrictions placed on age, gender, or diagnosis (or lack thereof). Self-harm can refer to aone-off or recurrent behaviour.

Concept: Interventions considered will exclusively focus on self-harm without an intention to end one's life. Any interventions for self-injurious behaviour where the intention was to die will be excluded. Collaborative involvement concepts vary in the literature with subtle distinctions between co-design, co-production, and co-creation. Co-design involves collaboration between parties to design a solution to a problem; co-production refers to collaboration post-design at the point of implementation; and co-creation is an overarchingterm referring to collaborative understanding of problems, as well as designing and evaluating solutions (Vargas et al., 2022). All terms will be included. Patient and public involvement (PPI) is a broader term that means working in partnership with the patient or public through all stages of research. As such the extent of involvement can vary and has

been classified along two axes: active-passive and embedded-isolated (Rethink Mental Illness, 2017). Papers must indicate an element of collaborative involvement of those with lived-experience in the title or abstract to be included. Literature will be excluded if involvement is restricted to consultation. Selecting all self-harm intervention literature andchecking the full text for evidence of co-design is beyond the scope of this review.

Context: The context for this review will be interventions across a broad range of locations including but not limited to primary, secondary, and tertiary care, digital, and self- management interventions. There will be no restriction on publication date in order to mapthe full extent of the self-harm co-design literature. The search will be conducted on 24$^{th}$ December 2022 so any literature published after this date will not be included.

Evidence sources:
This scoping review will consider primary studies, systematic reviews and meta analyses, aswell as conference abstracts, protocols, policy documents, guidelines and dissertations.
Primary sources will be excluded if already identified as part of an included review. Only English language studies will be included. There will be no limitations placed on the date ofevidence publication in order to capture the breadth of available evidence.

Search strategy:
An initial PubMed search was undertaken using ‘co-design’, ‘self-harm’, and ‘intervention’ to identify synonyms and related terms in the titles and abstracts of relevant articles. The following databases will be searched using the comprehensive search strategy developed: PubMed, Embase, PsycINFO, Web of Science and Cochrane Library, as well as grey literaturein PROSPERO and Clinical Trial and Results Registers. Department of Health, National Institute for Health and Care Research (NIHR), National Institute for Health and Care Excellence (NICE), and mental health charity websites such as The McPin Foundation will also be scoped for any relevant content. Backward and forward reference searches will alsobe conducted.

Due to the variety of terms used to describe both co-design and self-harm, a range of searchterms will be used (see Table 1). MeSH terms will be used in PubMed searches. If further relevant terms or sources of evidence become evident during the scoping process, these willbe scoped and documented. Duplicate articles will be removed prior to screening. Rayyan (https://www.rayyan.ai/) will be used to record decisions. All records will be screened for eligibility at title and abstract level, then at full-text level by two researchers independently (LCW and NLC). Any disagreements will be resolved via discussion to reach a consensus regarding inclusion.

Data extraction and presentation:
For each of the included full-text articles, key information will be extracted and charted by the primary author LCW (see Table 2). Data to be extracted may change during the scopingprocess thus any modifications will be recorded. Results of the scoping review will be arranged into themes to map the field and meet the objectives. Any gaps in the literature will be highlighted. This scoping review will be submitted as a medical student dissertation,and to a peer-reviewed journal for dissemination.

Table 1: search terms

| | | | | |
|---|---|---|---|---|
| co-design*<br>OR<br>codesign*<br>OR<br>co-prod*<br>OR<br>coprod*<br>OR<br>co-creat*<br>OR<br>cocreat*<br>OR<br>co-deliv*<br>OR<br>codeliv*<br>OR<br>co-evaluat*<br>OR<br>coevaluat*<br>OR<br>co-innov*<br>OR<br>coinnov*<br>OR<br>EBCD<br>OR<br>“patient and public involvement”<br>OR<br>PPI<br>OR<br>lived-experienc*<br>OR<br>“personal experienc*”<br>OR<br>participatory design* | AND | self-harm*<br>OR<br>self-injur*<br>OR<br>NSSI<br>OR<br>self-mutilat*<br>OR<br>DSH<br>OR<br>parasuicid*<br>OR<br>self-destructiv*<br>OR<br>self-cutting<br>OR<br>self-poiso*<br>OR<br>self-hitting<br>OR<br>self-inflict*<br>OR<br>head-bang*<br>OR<br>("Self-Injurious Behavior"[Mesh])<br>OR<br>("Self Mutilation"[Mesh]) | AND | intervention*<br>OR<br>approach*<br>OR<br>service*<br>OR<br>program*<br>OR<br>practice*<br>OR<br>manage*<br>OR<br>treatment*<br>OR<br>therap*<br>OR<br>prevent*<br>OR<br>resource*<br>OR<br>assessment* |

| |
|---|
| OR<br>participatory<br>research*<br>OR<br>“peer research*”<br>OR<br>LEAP |

| |
|---|
| OR<br>collaborative<br>involvement<br>OR<br>“experts by<br>experience”<br>OR<br>“expert by<br>experience”<br>OR<br>“service use*”<br>OR<br>“patient participation”<br>OR<br>“community<br>participation”<br>OR<br>stakeholder*<br>OR<br>("Community-Based<br>Participatory<br>Research"[Mesh])<br>OR<br>("Patient<br>Participation"[Mesh])<br>OR<br>("Community<br>Participation"[Mesh]) |

Table 2: Data to be extracted and charted from full-text articles:

- Citation
- Title
- Author(s)
- Year of publication
- Country of origin (where study was published/conducted)
- Type of publication (e.g. journal, guideline, protocol, dissertation)
- Study design (e.g. quantitative, qualitative, mixed method)
- Aims and objectives
- Methods
- Population (e.g. first-hand experience or carer, age range, diagnoses, gender, sample size)
- Setting (e.g. primary care, emergency department, online intervention)

- Self-harm (e.g. definition and method)
- Intervention
- Extent of co-design involvement (e.g. active, passive, embedded, isolated, consultation, decision-making, designing, authorship)
- Co-design benefits and challenges
- Co-design facilitators and barriers
- Any other key findings that relate to the scoping review's objectives

**Supplementary material 2. Complete search strategy**

| | | | | |
|---|---|---|---|---|
| co-design*<br>OR<br>codesign*<br>OR<br>co-prod*<br>OR<br>coprod*<br>OR<br>co-creat*<br>OR<br>cocreat*<br>OR<br>co-deliv*<br>OR<br>codeliv*<br>OR<br>co-evaluat*<br>OR | AND | self-harm*<br>OR<br>self-injur*<br>OR<br>NSSI<br>OR<br>self-mutilat*<br>OR<br>DSH<br>OR<br>parasuicid*<br>OR<br>self-destructiv*<br>OR<br>self-cutting<br>OR<br>self-poiso*<br>OR | AND | intervention*<br>OR<br>approach*<br>OR<br>service*<br>OR<br>program*<br>OR<br>practice*<br>OR<br>manage*<br>OR<br>treatment*<br>OR<br>therap*<br>OR<br>prevent*<br>OR |

| | | | | |
|---|---|---|---|---|
| coevaluat*<br>OR<br>co-innov*<br>OR<br>coinnov*<br>OR<br>EBCD<br>OR<br>“patient and public involvement”<br>OR<br>PPI<br>OR<br>lived-experienc*<br>OR<br>“personal experienc*”<br>OR<br>participatory design*<br>OR<br>participatory research*<br>OR<br>“peer research*”<br>OR<br>LEAP<br>OR<br>collaborative involvement<br>OR<br>“experts by experience”<br>OR<br>“expert by experience”<br>OR<br>“service use*”<br>OR<br>“patient participation”<br>OR<br>“community participation”<br>OR<br>stakeholder*<br>OR<br>("Community-Based Participatory Research"[Mesh]) | | self-hitting<br>OR<br>self-inflict*<br>OR<br>head-bang*<br>OR<br>("Self-Injurious Behavior"[Mesh])<br>OR<br>("Self Mutilation"[Mesh]) | | resource*<br>OR<br>assessment* |

| |
|---|
| OR ("Patient Participation"[Mesh]) OR ("Community Participation"[Mesh]) |

### Supplementary 3: example search using Pubmed

Search on Pubmed 24th December 2022

| Search number | Query | Sort By | Filters | Search Details | Results | Time |
|---|---|---|---|---|---|---|
| 8 | ((((co-design*[Title/Abstract] OR codesign*[Title/Abstract] OR co-prod*[Title/Abstract] OR coprod*[Title/Abstract] OR co-creat*[Title/Abstract] OR cocreat*[Title/Abstract] OR co-deliv*[Title/Abstract] OR codeliv*[Title/Abstract] OR co-evaluat*[Title/Abstract] OR coevaluat*[Title/Abstract] OR co-innov*[Title/Abstract] OR coinnov*[Title/Abstract] OR EBCD[Title/Abstract] OR "patient and public involvement"[Title/Abstract] OR PPI[Title/Abstract] OR lived-experienc*[Title/Abstract] OR "personal experienc*"[Title/Abstract] OR participatory design*[Title/Abstract] OR participatory research*[Title/Abstract] OR "peer research*"[Title/Abstract] OR LEAP[Title/Abstract] OR collaborative involvement[Title/Abstract] OR "experts by experience"[Title/Abstract] OR "expert by experience"[Title/Abstract] OR "service use*"[Title/Abstract] OR "patient participation"[Title/Abstract] OR "community participation"[Title/Abstract] OR stakeholder*[Title/Abstract])) OR (("Community-Based Participatory Research"[Mesh]) OR ("Patient Participation"[Mesh]) OR ("Community Participation"[Mesh]))) AND (((self-harm*[Title/Abstract] OR self-injur*[Title/Abstract] OR NSSI[Title/Abstract] OR self-mutilat*[Title/Abstract] OR DSH[Title/Abstract] OR parasuicid*[Title/Abstract] OR self-destructiv*[Title/Abstract] OR self-cutting[Title/Abstract] OR self-poiso*[Title/Abstract] OR self-hitting[Title/Abstract] OR self-inflict*[Title/Abstract] OR head-bang*[Title/Abstract])) OR (("Self- | | | ("co design*"[Title/Abstract] OR "codesign*"[Title/Abstract] OR "co prod*"[Title/Abstract] OR "coprod*"[Title/Abstract] OR "co creat*"[Title/Abstract] OR "cocreat*"[Title/Abstract] OR "co deliv*"[Title/Abstract] OR "codeliv*"[Title/Abstract] OR "co evaluat*"[Title/Abstract] OR "coevaluat*"[Title/Abstract] OR "co innov*"[Title/Abstract] OR "coinnov*"[Title/Abstract] OR "EBCD"[Title/Abstract] OR "patient and public involvement"[Title/Abstract] OR "PPI"[Title/Abstract] OR "lived experienc*"[Title/Abstract] OR "personal experienc*"[Title/Abstract] OR "participatory design*"[Title/Abstract] OR "participatory research*"[Title/Abstract] OR "peer research*"[Title/Abstract] OR "LEAP"[Title/Abstract] OR "collaborative involvement"[Title/Abstract] OR "experts by experience"[Title/Abstract] OR "expert by experience"[Title/Abstract] OR "service use*"[Title/Abstract] OR "Patient Participation"[Title/Abstract] OR "Community Participation"[Title/Abstract] OR "stakeholder*"[Title/Abstract] OR ("Community-Based Participatory Research"[MeSH Terms] OR "Patient Participation"[MeSH Terms] OR "Community Participation"[MeSH Terms])) AND ("self harm*"[Title/Abstract] OR "self injur*"[Title/Abstract] OR "NSSI"[Title/Abstract] OR "self mutilat*"[Title/Abstract] OR "DSH"[Title/Abstract] OR "parasuicid*"[Title/Abstract] OR "self destructiv*"[Title/Abstract] OR "self-cutting"[Title/Abstract] OR "self poiso*"[Title/Abstract] OR "self-hitting"[Title/Abstract] OR "self inflict*"[Title/Abstract] OR "head bang*"[Title/Abstract] OR ("Self-Injurious Behavior"[MeSH Terms] OR "Self | 1,122 | 16:03:43 |

| | | | | | | |
|---|---|---|---|---|---|---|
| | Injurious Behavior"[Mesh]) OR ("Self Mutilation"[Mesh])))) AND ((intervention*[Title/Abstract] OR approach*[Title/Abstract] OR service*[Title/Abstract] OR program*[Title/Abstract] OR practice*[Title/Abstract] OR manage*[Title/Abstract] OR treatment*[Title/Abstract] OR therap*[Title/Abstract] OR prevent*[Title/Abstract] OR resource*[Title/Abstract] OR assessment*[Title/Abstract])) | | | Mutilation"[MeSH Terms])) AND ("intervention*"[Title/Abstract] OR "approach*"[Title/Abstract] OR "service*"[Title/Abstract] OR "program*"[Title/Abstract] OR "practice*"[Title/Abstract] OR "manage*"[Title/Abstract] OR "treatment*"[Title/Abstract] OR "therap*"[Title/Abstract] OR "prevent*"[Title/Abstract] OR "resource*"[Title/Abstract] OR "assessment*"[Title/Abstract]) | | |
| **7** | (intervention*[Title/Abstract] OR approach*[Title/Abstract] OR service*[Title/Abstract] OR program*[Title/Abstract] OR practice*[Title/Abstract] OR manage*[Title/Abstract] OR treatment*[Title/Abstract] OR therap*[Title/Abstract] OR prevent*[Title/Abstract] OR resource*[Title/Abstract] OR assessment*[Title/Abstract]) | | | "intervention*"[Title/Abstract] OR "approach*"[Title/Abstract] OR "service*"[Title/Abstract] OR "program*"[Title/Abstract] OR "practice*"[Title/Abstract] OR "manage*"[Title/Abstract] OR "treatment*"[Title/Abstract] OR "therap*"[Title/Abstract] OR "prevent*"[Title/Abstract] OR "resource*"[Title/Abstract] OR "assessment*"[Title/Abstract] | 12,680,553 | 15:58:27 |
| **6** | ((self-harm*[Title/Abstract] OR self-injur*[Title/Abstract] OR NSSI[Title/Abstract] OR self-mutilat*[Title/Abstract] OR DSH[Title/Abstract] OR parasuicid*[Title/Abstract] OR self-destructiv*[Title/Abstract] OR self-cutting[Title/Abstract] OR self-poiso*[Title/Abstract] OR self-hitting[Title/Abstract] OR self-inflict*[Title/Abstract] OR head-bang*[Title/Abstract])) OR (("Self-Injurious Behavior"[Mesh]) OR ("Self Mutilation"[Mesh])) | | | "self harm*"[Title/Abstract] OR "self injur*"[Title/Abstract] OR "NSSI"[Title/Abstract] OR "self mutilat*"[Title/Abstract] OR "DSH"[Title/Abstract] OR "parasuicid*"[Title/Abstract] OR "self destructiv*"[Title/Abstract] OR "self-cutting"[Title/Abstract] OR "self poiso*"[Title/Abstract] OR "self-hitting"[Title/Abstract] OR "self inflict*"[Title/Abstract] OR "head bang*"[Title/Abstract] OR "Self-Injurious Behavior"[MeSH Terms] OR "Self Mutilation"[MeSH Terms] | 91,039 | 15:58:17 |
| **5** | ("Self-Injurious Behavior"[Mesh]) OR ("Self Mutilation"[Mesh]) | | | "Self-Injurious Behavior"[MeSH Terms] OR "Self Mutilation"[MeSH Terms] | 81,570 | 15:58:12 |
| **4** | (self-harm*[Title/Abstract] OR self-injur*[Title/Abstract] OR NSSI[Title/Abstract] OR self-mutilat*[Title/Abstract] OR DSH[Title/Abstract] OR parasuicid*[Title/Abstract] OR self-destructiv*[Title/Abstract] OR self-cutting[Title/Abstract] OR self-poiso*[Title/Abstract] OR self-hitting[Title/Abstract] OR self-inflict*[Title/Abstract] OR head-bang*[Title/Abstract]) | | | "self harm*"[Title/Abstract] OR "self injur*"[Title/Abstract] OR "NSSI"[Title/Abstract] OR "self mutilat*"[Title/Abstract] OR "DSH"[Title/Abstract] OR "parasuicid*"[Title/Abstract] OR "self destructiv*"[Title/Abstract] OR "self-cutting"[Title/Abstract] OR "self poiso*"[Title/Abstract] OR "self-hitting"[Title/Abstract] OR "self inflict*"[Title/Abstract] OR "head bang*"[Title/Abstract] | 21,687 | 15:57:59 |
| **3** | ((co-design*[Title/Abstract] OR codesign*[Title/Abstract] OR co-prod*[Title/Abstract] OR coprod*[Title/Abstract] OR co-creat*[Title/Abstract] OR cocreat*[Title/Abstract] OR co-deliv*[Title/Abstract] OR codeliv*[Title/Abstract] OR co-evaluat*[Title/Abstract] OR coevaluat*[Title/Abstract] OR co-innov*[Title/Abstract] OR | | | "co design*"[Title/Abstract] OR "codesign*"[Title/Abstract] OR "co prod*"[Title/Abstract] OR "coprod*"[Title/Abstract] OR "co creat*"[Title/Abstract] OR "cocreat*"[Title/Abstract] OR "co deliv*"[Title/Abstract] OR "codeliv*"[Title/Abstract] OR "co evaluat*"[Title/Abstract] OR "coevaluat*"[Title/Abstract] OR "co innov*"[Title/Abstract] OR | 200,008 | 15:57:35 |

| | | | | | | |
|---|---|---|---|---|---|---|
| | coinnov*[Title/Abstract] OR EBCD[Title/Abstract] OR "patient and public involvement"[Title/Abstract] OR PPI[Title/Abstract] OR lived-experienc*[Title/Abstract] OR "personal experienc*"[Title/Abstract] OR participatory design*[Title/Abstract] OR participatory research*[Title/Abstract] OR "peer research*"[Title/Abstract] OR LEAP[Title/Abstract] OR collaborative involvement[Title/Abstract] OR "experts by experience"[Title/Abstract] OR "expert by experience"[Title/Abstract] OR "service use*"[Title/Abstract] OR "patient participation"[Title/Abstract] OR "community participation"[Title/Abstract] OR stakeholder*[Title/Abstract])) OR (("Community-Based Participatory Research"[Mesh]) OR ("Patient Participation"[Mesh]) OR ("Community Participation"[Mesh])) | | | "coinnov*"[Title/Abstract] OR "EBCD"[Title/Abstract] OR "patient and public involvement"[Title/Abstract] OR "PPI"[Title/Abstract] OR "lived experienc*"[Title/Abstract] OR "personal experienc*"[Title/Abstract] OR "participatory design*"[Title/Abstract] OR "participatory research*"[Title/Abstract] OR "peer research*"[Title/Abstract] OR "LEAP"[Title/Abstract] OR "collaborative involvement"[Title/Abstract] OR "experts by experience"[Title/Abstract] OR "expert by experience"[Title/Abstract] OR "service use*"[Title/Abstract] OR "Patient Participation"[Title/Abstract] OR "Community Participation"[Title/Abstract] OR "stakeholder*"[Title/Abstract] OR "Community-Based Participatory Research"[MeSH Terms] OR "Patient Participation"[MeSH Terms] OR "Community Participation"[MeSH Terms] | | |
| **2** | ("Community-Based Participatory Research"[Mesh]) OR ("Patient Participation"[Mesh]) OR ("Community Participation"[Mesh]) | | | "Community-Based Participatory Research"[MeSH Terms] OR "Patient Participation"[MeSH Terms] OR "Community Participation"[MeSH Terms] | 52,301 | 15:57:28 |
| **1** | (co-design*[Title/Abstract] OR codesign*[Title/Abstract] OR co-prod*[Title/Abstract] OR coprod*[Title/Abstract] OR co-creat*[Title/Abstract] OR cocreat*[Title/Abstract] OR co-deliv*[Title/Abstract] OR codeliv*[Title/Abstract] OR co-evaluat*[Title/Abstract] OR coevaluat*[Title/Abstract] OR co-innov*[Title/Abstract] OR coinnov*[Title/Abstract] OR EBCD[Title/Abstract] OR "patient and public involvement"[Title/Abstract] OR PPI[Title/Abstract] OR lived-experienc*[Title/Abstract] OR "personal experienc*"[Title/Abstract] OR participatory design*[Title/Abstract] OR participatory research*[Title/Abstract] OR "peer research*"[Title/Abstract] OR LEAP[Title/Abstract] OR collaborative involvement[Title/Abstract] OR "experts by experience"[Title/Abstract] OR "expert by experience"[Title/Abstract] OR "service use*"[Title/Abstract] OR "patient participation"[Title/Abstract] OR "community participation"[Title/Abstract] OR stakeholder*[Title/Abstract]) | | | "co design*"[Title/Abstract] OR "codesign*"[Title/Abstract] OR "co prod*"[Title/Abstract] OR "coprod*"[Title/Abstract] OR "co creat*"[Title/Abstract] OR "cocreat*"[Title/Abstract] OR "co deliv*"[Title/Abstract] OR "codeliv*"[Title/Abstract] OR "co evaluat*"[Title/Abstract] OR "coevaluat*"[Title/Abstract] OR "co innov*"[Title/Abstract] OR "coinnov*"[Title/Abstract] OR "EBCD"[Title/Abstract] OR "patient and public involvement"[Title/Abstract] OR "PPI"[Title/Abstract] OR "lived experienc*"[Title/Abstract] OR "personal experienc*"[Title/Abstract] OR "participatory design*"[Title/Abstract] OR "participatory research*"[Title/Abstract] OR "peer research*"[Title/Abstract] OR "LEAP"[Title/Abstract] OR "collaborative involvement"[Title/Abstract] OR "experts by experience"[Title/Abstract] OR "expert by experience"[Title/Abstract] OR "service use*"[Title/Abstract] OR "patient participation"[Title/Abstract] OR "community participation"[Title/Abstract] OR "stakeholder*"[Title/Abstract] | 156,957 | 15:57:12 |
